# Break-through COVID-19 infection rate with Indian strain in Single-center Healthcare Workers – A real world data

**DOI:** 10.1101/2021.07.02.21258881

**Authors:** Ravindra Sabnis, Abhijit Patil, Nitiraj Shete, Arun Kumar Rastogi

## Abstract

**Introduction:** It is observed that many healthcare workers got COVID-19 infection despite of completing both doses of Covishield vaccine. This study aimed to find real incidence of vaccine breakthrough infection.

**Material and methods:** All hospital employees, who were fully vaccinated were included in study. Details about their vaccine side effects, infection prior to vaccination, post vaccination infection, severity of infection, hospital and ICU admission were noted.

**Results:** None encountered any significant side effects of vaccine. Of the 461 participants – 86 (18.65%) got infection average 38 days (range 14-70days) after vaccination. As per the NIH classification, out of 86, disease was mild in 69(80.2%), moderate in 10(11.62%), severe in 6(6.97%) and critical in 1(1.16%). Of these, 10(11.62%) required hospital admission. Of these 10, 2 were shifted to ICU. Of the 2, One recovered while one died. Thus mortality was 1/86(1.6%).

**Conclusion:** Breakthrough infection rate in health care workers was 18.65%. Moderate, severe or critical disease occurred in 19.7% participants even after two doses of vaccine. Mortality due to disease cannot be completely obviated due to vaccine. The vaccine was safe without any significant adverse events.

## Introduction

COVID-19 infection which is caused by severe acute respiratory syndrome coronavirus 2 (SARS-CoV-2) had first confirmed case at Wuhan, China at December 2019. WHO declared COVID-19 pandemic spread across the world in March 2020. As of 10 June 2021, more than 174 million cases have been confirmed, with more than 3.75 million confirmed deaths attributed to COVID-19, making it one of the deadliest pandemics in history.(1) This stimulated rapid development of vaccine against virus by many companies. AstraZeneca developed ChAdOx1 nCov-19 (AZD1222) vaccine. A safety and efficacy in randomized controlled trial of this vaccine performed in Brazil, South Africa & UK found the vaccine to have acceptable safety & efficacious against symptomatic COVID-19 infection.(2) In India same vaccine was licensed and manufactured by Serum Institute of India Private Limited(Pune, India) under the name of Covishield (AstraZeneca-Oxford) which was available for clinical use from mid-January 2021. All healthcare workers were given priority in first stage of vaccination program in India. Accordingly, the vaccination of all healthcare workers was started at our institute at end of January 2021 with second dose after a gap of 4 weeks.

The second wave of COVID-19 infection hit India in mid of March. Within short time, infection spread very rapidly. It was thought to be different strain – Indian strain – B.1.617(3) According to WHO, any variant which increases transmissibility, virulence of disease and decreases the efficacy of vaccine is known as variant of concern. The Indian-Delta variant (B.1.617.2) is designated such variant of concern.(4) In second wave, several healthcare workers got infection in spite of taking both doses of vaccine. Some of them had moderate to severe disease requiring hospital /ICU admission & even death. In the safety and efficacy trial conducted by AstraZeneca, it was found that vaccine was very effective in preventing symptomatic infection, hospital/ICU admissions and offered 100% protection from death.(2) The real world experience of the vaccine is quite different than the trial population.(5) Hence, we decided to conduct study with primary objective of finding out breakthrough infection rate among healthcare workers. Secondary objectives were to determine the severity of infection requiring hospital / ICU admissions or any mortality and the safety of vaccine.

## Material methods

This was a prospective study involving all the health care workers at a tertiary health care center. All the health care workers working at our institute were vaccinated with two doses of Covishield (SII-ChAdOx1 nCoV-19) administered on Days 1 and 29 as 0.5 ml dose intramuscularly. As a hospital policy, hospital maintained the health-related data from COVID-19 point of view of all employees after vaccination. If any information was lacking or more details were required, that employee was contacted to obtain it. The institutional ethical approval was achieved.

All the healthcare workers more than age 18 years who voluntarily consented for vaccine and data analysis were included in the study. The health-related data was retrieved from the hospital registry after consent of the participant. The data was collected from first dose of COVID-19 vaccine till 10^th^ June 2021. The data included the basic and the clinical data demography and family history of the participants.

The severity of disease was categorised as per the NIH guidelines of COVID-19 treatment (6)

### Asymptomatic or Presymptomatic Infection

Individuals who test positive for SARS-CoV-2 using a virologic test (i.e., a nucleic acid amplification test [NAAT]) but who have no symptoms that are consistent with COVID-19.

### Mild Illness

Individuals who have any of the various signs and symptoms of COVID-19 (e.g., fever, cough, sore throat, malaise, headache, muscle pain, nausea, vomiting, diarrhoea, loss of taste and smell) but who do not have shortness of breath, dyspnoea, or abnormal chest imaging.

### Moderate Illness

Individuals who show evidence of lower respiratory disease during clinical assessment or imaging and who have an oxygen saturation (SpO2) ≥94% on room air at sea level.

### Severe Illness

Individuals who have SpO2 <94% on room air at sea level, a ratio of arterial partial pressure of oxygen to fraction of inspired oxygen (PaO2/FiO2) <300 mm Hg, respiratory frequency >30 breaths/min, or lung infiltrates >50%.

### Critical Illness

Individuals who have respiratory failure, septic shock, and/or multiple organ dysfunction.

The statistical analysis was done using software IBM SPSS version 25 (IBM, Armonk, NY, USA). Quantitative variables were described as mean±SD while qualitative variables were described as %. Significance of qualitative variables were tested using proportion test. p-value of less than 0.05 was considered to be significant.

## Results

There are 506 total hospital employees, of which 19 were not vaccinated because of various stages of pregnancy & lactation while 26 did not take vaccine voluntarily. So total 461 employees completed both the doses of vaccine and qualified to be included in study. All 461 employees consented to participate in project and so the participation rate was 100%. There were 51 doctors in the cohort.

### Adverse events after vaccination

277/461(60.08%) participants were asymptomatic while the remaining 184/461(39.92%) participants were symptomatic after vaccination. Of the symptomatic cohort, 106/184(23%) had symptoms after first dose, 26/184(5.64%) had symptoms after second dose and 52/184(11.28%) had symptoms after both the doses. Of 184 symptomatic cohort, 96 (52.17%) did not require any treatment, 75 (40.76%) had mild symptoms requiring only paracetamol for < 2days. 13 (7.07%) had mild symptoms which lasted for > 2 days. None had any major adverse events after vaccine.

### Breakthrough COVID-19 infection after vaccination

86/461 (18.65%) had RTPCR proved COVID-19 symptomatic infection post-vaccination. The mean duration of infection from second dose of vaccine was 38.42±12.2 days (range 17-70 days). No participant had infection between first and second dose of vaccination. The mean age of the breakthrough infection cohort was 34.18 years(range – 22-77 years). Of these health-care workers, 19/86 (22.09%) were doctors, 27/86 (31.40%) were nurses/technicians and 40/86 (46.5%) were para-medical staff. 9/86 (10.6%) participants had co-morbidities (Diabetes milletus in 4, hypertension in 6, ischemic heart disease in 1).

### Severity of breakthrough infection

The infection was mild in 69/86 (80.2%), moderate in 10/86 (11.62%), severe in 6/86 (6.97%) and critical in 1/86 (1.16%). Of these, 10 (11.62%) required hospital admission. The indications for hospital admissions were – prolonged lower respiratory symptoms, significant lung involvement with blood abnormalities, requirement of IV medications, dyspnoea or oxygen saturation falling below 94. Of these 10 hospitalized patients, 2 had to be shifted to ICU for high flow oxygen & respiratory failure. Of these 2, one recovered and the other required ventilatory support due to falling saturation and despite all the efforts, this patient died. Thus, mortality rate in our study population was 1/86 (1.16%) - this was a healthcare worker with only hypertension as comorbidity and symptoms of cough, body ache, malaise after 31 days of second dose of vaccination. Later developed dyspnoea on walking & had blood abnormalities. Initially was managed conservatively with oral medications. But needed hospitalization for IV steroids and oxygen supplementation for dyspnoea. Patient was shifted to intensive care unit and required ventilatory support, but died on 7^th^ day of infection. There was severe disease in 7/86 (8.13%) patients requiring oxygen supplementation for variable period of time from 12 hrs to 72 hrs.

All the family members of two healthcare workers were infected with COVID-19. None of these family members were vaccinated. Both the healthcare workers being fully vaccinated had mild disease course. On the contrary, all the affected family members had severe disease with one death of family member in each family.

### Laboratory abnormalities

Blood parameters were done in 48/86 (55.81%). Among these 48 participants, C-reactive protein(CRP) was raised (Cut off value – 7mg/L) in 17 cases. Mean CRP was 23.4 mg/L (range 10-73). d-DIMER (Lab cut off - 0.5 mcg/ml) was deranged in 26 cases. Mean d-DIMER was 0.89 mcg/ml(range 0.6 - 3.1 mcg/ml). Chest CT scan was performed in 21 participants, which was abnormal in 18 participants (CT severity score – 5 to 16/25).

### Pre-vaccination COVID-19 infection

40 participants had infection at least two months prior to first dose of vaccination. The mean age was 39.9±12years(range 23-68 years). 4/40(10%) were doctors, 17/40(42.5%) were nurses or technicians while 19/40(47.5%) were paramedical staff. The infection was mild in 25(62.5%), moderate in 11 (27.5%) and severe in 4 (10%) cases. 7 (17.5%) required hospital admission of which one needed ICU care. Oxygen support was needed in 4 cases. All recovered from treatment. There was no mortality in this group. All these patients took vaccine after they recovered.

### Reinfection in post-vaccination period

Out of these 40 previously infected participants, 10(25%) had break-through infection. All had received both doses of vaccine. All had mild disease and recovered without any specific treatment.

The family members or close contact group were infected in 9/86 (10.46%) post-vaccination cohort. However, this infection rate was 11/40(27.5%) in the pre-vaccination era.

## Discussion

All eligible hospital employees participated in the study. Thus, removing attrition bias. All the healthcare workers were always available for health update and thus their data was accurate, obviating recall bias. This was the strength of the study.

Vaccination program started at end of January 2021 at our hospital. The incidence of COVID-19 was low at our local region along with whole country. The second wave started at mid March 2021 during which the incidence of COVID-19 infected patients increased at our hospital. By this time all the health care workers had finished both the doses of vaccine, hence none in our study got infection between 1^st^ and 2^nd^ dose of vaccine as their exposure to virus was very low. First infection happened 14 days after second dose and last was after 70 days, average being 38 days. Studies have shown adequate seroconversion by 2 weeks.(7). Thus it is assumed that all healthcare workers had adequate immunity at the time of breakthrough infection.

In our study, symptomatic infection rate after both doses of vaccination was 18.65%. The average vaccine efficacy reported in an interim analysis of four randomised controlled trials in Brazil, South Africa, and the UK was around 70.4%.(2) In a further pooled analysis of previous four trials on this vaccine, overall vaccine efficacy more than 14 days after the second dose was 66·7% and requiring no hospital admission. (8) The AstraZeneca US Phase III trial of AZD1222 interim safety and efficacy analysis based on 32,449 participants accruing 141 symptomatic cases of COVID-19 demonstrated vaccine efficacy of 79% at preventing symptomatic COVID-19.(9) A study found decreased efficacy of this vaccine against the delta variant 66.1% to 59.8%. (10) The efficacy of vaccine in our study against symptomatic disease was 81.3%.

In our study, mild disease was seen in 80.2%, while it was either moderate, severe or critical disease as per NIH criteria in 19.8%. Out of 86, 10 (11.62%) required hospital admission, of which 2 had to be shifted to ICU. This is contrary to no patients requiring hospitalisation (WHO clinical progression score ≥4) and no patients with Severe COVID-19 (WHO clinical progression score ≥6) in the trials on Covishield vaccine.(2,9) This higher percentage may be because of Indian strain B.1.617.2(Delta variant) which is shown to be more virulent.(11) There was no death in the largest trial assessing the efficacy of this vaccine.(2) However in our study, 1/86 (1.16%) death was encountered, who was in relatively younger age group with only hypertension as co-morbidities. This again could be attributed to virulent strain or genetic susceptibility of person to COVID-19 infection.

All the family members of two healthcare workers were infected with COVID-19. None of these family members were vaccinated. Both the healthcare workers being fully vaccinated had moderate disease course. On the contrary, all the affected family members had severe disease with one death of family member in each family. Although genetic susceptibility of a person to COVID-19 infection is not proved fact, there is possibility of infection in family members with similar genetic composition in everybody. This is only observation which requires further evaluation.

40 participants had previous infection. Doctors constituted only 4/51(7.8%) of this cohort during first wave, while in second wave, doctors constituted 19/51(37.25%). This indicates that doctors were affected in large number during second wave (p=0.001). The probable reason for this could be high level of exposure to their treating patient with COVID-19 infection and increased virulence of virus. Moderate, severe or critical disease was present in 37.5% in pre-vaccination group while only 19.7% in post vaccination infection group. This was statistically significant (p=0.043). This reduced severity may be due to the vaccine. However, there was no mortality in pre-vaccination group, while there was one death in post vaccination cohort. 10 participants had breakthrough COVID-19 infection after previous infection in pre-vaccination period. However, in all cases, disease was mild. Reinfection suggests some different genetic composition of virus, against which antibodies were not present either due to previous infection or vaccination. This supports the report that original virus has undergone mutations namely L452R, E484Q and P681R.(3).

In our study, family members and close contacts of 10.46% participants were infected in post-vaccination group as against 27.5% infection rate of family members and close contacts in pre-vaccination group. (p=0.029) This suggests that spread from vaccinated people to others is significantly less. This finding is similar to other studies which showed that spreading potential post vaccination was much less.(12,13)

Several studies have proved safety profile of vaccine in trial population.(2,14,15) In our study, none had any significant side effects of vaccine, early or late, supporting that vaccine is safe even in real world scenario.

## Conclusion

The post-vaccine infection rate in health care workers was 18.65%. Moderate, severe or critical disease occurred in 19.7% participants even after two doses of vaccine. Mortality due to disease cannot be completely obviated due to vaccine. The vaccine was safe without any significant adverse events.

## Supporting information

Ethical approval file

## Data Availability

NA

## Author contributions

**R.B.S**. : Concept, Data collection and manuscript supervision

**A.P**. : Data collection, data analysis and manuscript writing

**N.S**. : Data analysis

**A.K.R**. : Data collection supervision

## Competing interests

None

## Funding source

None

---

**Table 1:**
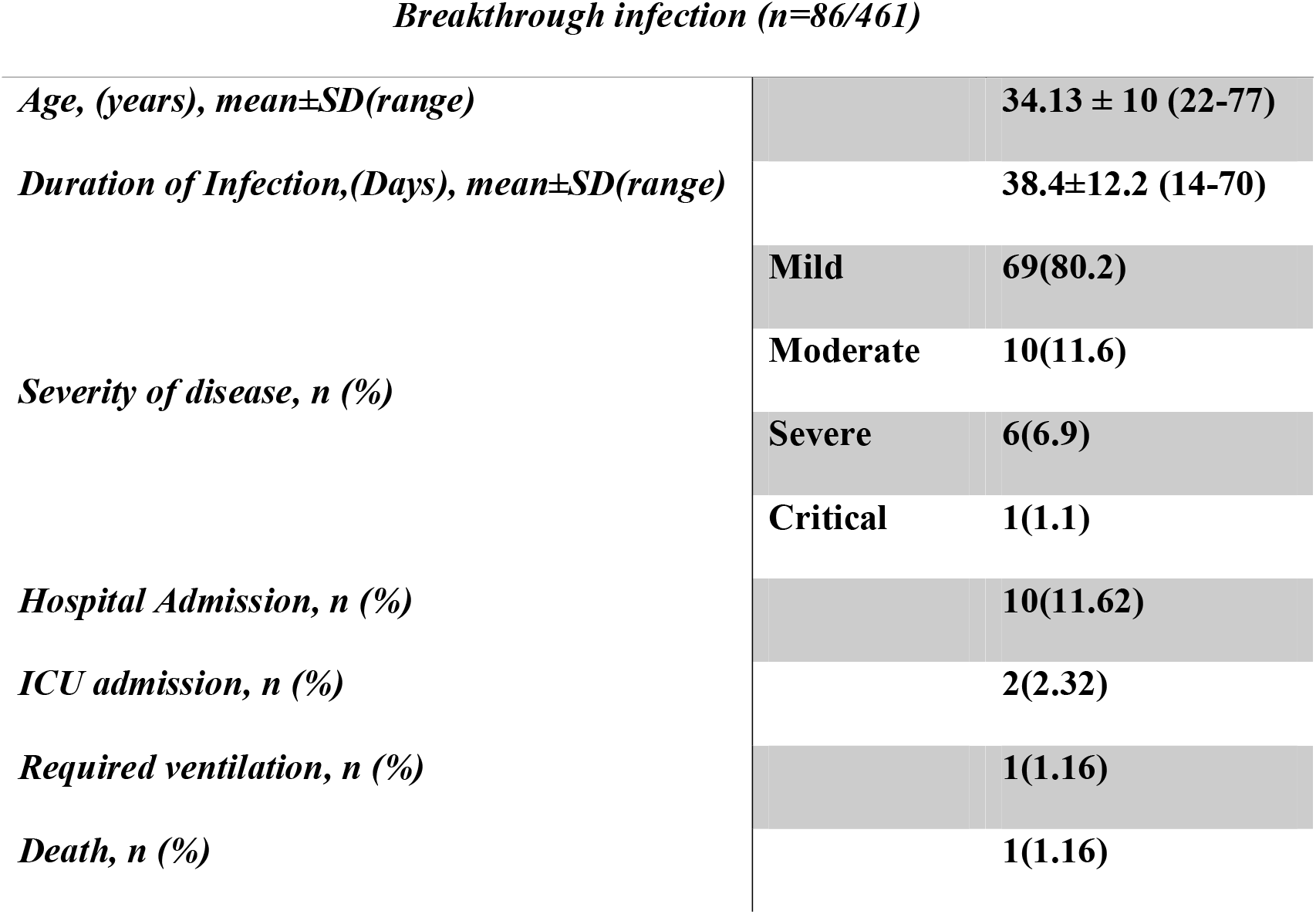
Details of break-through infection.

## Notes

### Competing Interest Statement

The authors have declared no competing interest.

### Clinical Trial

CTRI/2021/06/034019

### Funding Statement

No funding or conflict of interest.

### Author Declarations

Full name of the ethics committee: Muljibhai Patel Society for Research in Nephro-Urology Ethics committee

Institutional affiliation of the ethics committee: Muljibhai Patel Urological Hospital, Nadiad, Gujarat, India

Decision of the ethics committee: Muljibhai Patel Society for Research in Nephro-Urology Ethics committee approve the trial to be conducted in its presented form.

