## Supplementary material for "Break-through COVID-19 infection rate with Indian strain in Single-center Healthcare Workers – A real world data": Ethical approval file

### Muljibhai Patel Society For Research in Nephro - Urology

Dr. VIRENDRA DESAI ROAD, NADIAD - 387 001.
 Website : www.mpuh.org

Date: 03rd May 2021

Dr. Ravindra Sabnis
Consultant Urologist,
Muljibhai Patel Urological Hospital
Dr. Virendra Desai Road
Nadiad 387001, Gujarat, India

Reference: EC/728/2021

Dear Dr. Ravindra Sabnis

The institutional ethics committee Muljibhai Patel Society for Research in Nephro-Urology Ethics committee reviewed and discussed your application to conduct the clinical trial entitled "**Breakthrough COVID-19 infection rate with Indian strain in Single-centre Healthcare workers- A real world data**" on 29th April 2021.

The following documents were reviewed:

1. Introduction
2. Methodology
3. Result

The following members of the Muljibhai Patel Society for Research in Nephro-Urology Ethics committee were present at the meeting held on 29th April 2021 at 03:30 pm on the Zoom video conference platform.

| Sr. No. | Name of the Member | Position in Ethics Committee | Designation & Affiliation | Qualification | Gender |
|---|---|---|---|---|---|
| 1 | Dr. Rucha Mehta | Chair person | Endocrinologist, Apollo Hospital, Ahmedabad | MD | Female |
| 2 | Dr. Minaxi Shah | Interim Chair person | Professor, Department of Pharmacology Gujarat Cancer Society Medical college, Hospital , Ahmedabad | M.B.B.S, MD | Female |
| 3 | Dr. Barna Ganguly | Pharmacologist | Professor of Department of Pharmacology and HEAD,UNESCO Bioethics Unit, Gujarat, India | M.D , PG Diploma Bioethics | Female |
| 4 | Dr. Mahesh Desai | Clinician | Managing Trustee, Department of Urology, Consultant Urologist, Muljibhai Patel Urological Hospital, Nadiad | MS,DM, DNB, | Male |
| 5 | Dr. Ravindra Sabnis | Clinician | Chairman, Department of Urology, Consultant Urologist, Muljibhai Patel Urological Hospital, Nadiad | M.B.B.S,M.S, M.Ch | Male |

| | | | | | |
|---|---|---|---|---|---|
| 6 | Dr. Pankaj Parikh | Clinician | General Surgeon, Sainath Hospital | M.B.B.S, M.S | Male |
| 7 | Dr. Sishir Gang | Clinician | Chairman, Department of Nephrology, Consultant Nephrologist, Muljibhai Patel Urological Hospital, Nadiad | MD,DM,DN | Male |
| 8 | Dr. Shailesh Soni | Member Secretary | Chief Pathologist, Muljibhai Patel Urological Hospital, Nadiad | MD | Male |
| 9 | Mrs. Hema Joshipura | Layman | Retired Banker | B.Com, LLB, cs | Female |
| 10 | Mr. Bipin Patel | Legal Expert | Advocate | B.Sc., L.L.B., D.L.P., D.P.M. (Bhavan),Dip. in Ind. pscy., D.H.R.D ,D.B.A., M.B.A.(HR | Male |
| 11 | Mrs. Minal Thakar | Social Scientist | Educationalist | M.A.Ed (English) | Female |

**Muljibhai Patel Society for Research in Nephro-Urology Ethics committee approve the trial to be conducted in its presented form.**

The Ethics Committee to be informed about the progress of the study, any Serious Adverse Events (SAE) occurring in the course of the study, any changes in the protocol and patient information or informed consent and to be provided with a copy of the final report. Yours sincerely,

03/05/2021

Dr. Shailesh Soni                                        Date: 03rd May 2021
Member Secretary
Muljibhai Patel Society for Research in Nephro-Urology Ethics Committee
Muljibhai Patel Urological hospital,
Dr. Virendra Desai Road,
Nadiad — 387001, Gujarat, India
